# Interrogating a framework for diabetic retinopathy screening adherence: qualitative interviews of a severe disease and unengaged population

**DOI:** 10.1101/2025.05.29.25328601

**Authors:** Julia Fu, Joana Andoh, Elizabeth Fairless, June Weiss, Althea Norcott, Kristen Nwanyanwu

**Author notes:** Corresponding author (KN).

## Abstract

**Background:** To interrogate a framework of diabetic retinopathy (DR) screening adherence by conducting qualitative interviews with individuals with severe DR and those unengaged in eye care.

**Methods:** From March 2021 to February 2022, we conducted eight remote semi-structured interviews with participants diagnosed with diabetes divided into two cohorts: those with severe DR who had undergone procedures (n=4) and those unengaged in eye care for >1 year (n=4). We recruited participants from an academic faculty practice, community referrals, and word of mouth. During the interviews, we collected demographic data and presented participants with a DR screening utilization framework previously developed by our group. We transcribed all interviews and conducted analyses using grounded theory and the constant comparative method to identify recurring themes.

**Results:** In the unengaged cohort, seven recurring themes emerged: *vision status*, *emotional context*, *competing concerns*, *resource availability*, *cues to action*, *knowledge-creating experiences*, *and in-clinic experiences*. These themes also emerged in the severe disease cohort, with the addition of the *patient-doctor relationship*. At the individual level, participants with stable vision often perceived no need for screening. At the interpersonal level, participants identified powerful patient-doctor relationships that empowered them to seek care. At the institutional and structural level, participants identified lack of insurance and transportation as significant barriers.

**Conclusions:** Improving education about DR, increasing resource accessibility, and strengthening the patient-doctor relationship may mitigate barriers to DR screening in populations at highest-risk for vision loss. To do so, we must implement innovative strategies, such as co-designed educational videos and digital health tools.

## Introduction

Diabetic retinopathy (DR) is a leading cause of blindness among working-age adults in the United States. Approximately 9.6 million people currently live with DR, and around 1.84 million experience vision-threatening DR [1]. Current guidelines recommend annual eye screening for individuals with diabetes, which may aid in early detection and prevention of vision loss [2–3]. However, many individuals are not screened in a timely fashion. It is estimated that only 56.9% of the more than 93 million US adults at high risk for vision loss receive eye care annually [4]. Therefore, it is crucial to understand the factors that influence DR screening adherence and develop relevant interventions to improve screening access.

Previous studies have explored DR screening adherence through qualitative methods, which enable a deeper understanding of peoples’ diverse experiences [5]. For example, Hartnett et. al. conducted focus groups with individuals with diabetes and identified key barriers to DR screening such as financial difficulties and limited access to services [6]. Similarly, Elam et. al. conducted focus groups with individuals at high risk for vision loss and found that common barriers to eye care included high costs, lack of trust in healthcare providers, clinic inaccessibility, and poor patient-doctor relationships [7].

Our research team previously developed a framework to understand DR screening adherence by conducting qualitative interviews with individuals with diabetes at high-risk for vision loss [8]. This framework outlined the individual and institutional/structural factors affecting DR screening adherence. Individual factors included vision status, competing concerns, and the emotional context of receiving screenings. Institutional/structural factors included resource availability, cues to action, knowledge-creating experiences, and in-clinic experiences.

In this study, we aim to validate and expand upon this framework developed in our previous study by conducting qualitative interviews of individuals with diabetes with severe disease and those not engaged in eye care. By including these new cohorts of individuals, we hope to gain a broader and more comprehensive understanding of the factors influencing DR screening adherence.

## Methods

The institutional review board (IRB) of Yale University approved this study. This study abided by the tenets of the Declaration of Helsinki. Leaders of local community health organizations through the Yale Community Engaged Research Steering Committee advised the study design.

Between March 9^th^, 2021, and February 17^th^, 2022, we recruited participants through the Yale Eye Center, via referrals from community partners, and by word of mouth. As part of recruitment, we contacted participants via phone or email to introduce the study and gauge their interest. The researchers had no prior relationships with the participants.

We conducted eight individual semi-structured, qualitative interviews in English with participants previously diagnosed with diabetes divided into two cohorts–severe disease and those unengaged in eye care. The severe disease cohort included individuals with DR who had undergone laser surgery or other procedures. The unengaged cohort included individuals with diabetes who had not received an eye exam in the past year. Each cohort consisted of four participants. We interviewed eight participants to reach thematic saturation.

We conducted interviews remotely by phone or web-based videoconferencing software (Zoom, San Jose, CA) and recorded interviews for transcription. Study author J.A. (female, medical student) conducted the interviews based on the principles of grounded theory. J.A. completed online training modules in qualitative research methods through Yale University and studied the textbook, *Qualitative Research: A Guide to Design and Implementation (*4th Edition, ISBN: 978-1-119-00361-8). Only participants and researchers were present during the interviews. Prior to each interview, we obtained verbal informed consent for participation in the study and documented consent via recording and transcription of the interview. We did not obtain written informed consent due to the remote nature of the interviews. The IRB of Yale University approved our use of verbal consent.

At the start of each interview, we introduced participants to the researcher, including her educational background and interest in the topic, and shared the purpose of the study. We then collected basic demographic information including age, gender, race, ethnicity, insurance status, and duration of diabetes. During the interview, we presented participants with a slide deck (S1 Appendix) containing the thematic framework previously developed by our research group, which explored the barriers and facilitators to DR screening adherence in a high-risk cohort [8]. The interview guide is found in Table 1. Each interview lasted approximately 60 minutes. After the interview, we mailed participants a $20 gift card. We did not carry out repeat interviews.

**Table 1.** Interview Guide.

|  |
| --- |
| **Resource Availability**<br>Has anything ever prevented you from having an eye exam?<br>Have you ever cancelled or not shown up to an appointment? If so, why?<br>Is there anything that would make it easier for you to get an eye exam?<br>Who or what has helped you stay on top of your eye exams?<br>If barrier X was removed, what do you think would happen? |
| **Cues to Action**<br>What made you decide to get/not get a dilated eye exam?<br>What brought you here today? What brought you to your last eye exam?<br>What factors motivated you? What factors reminded you?<br>Has anyone encouraged you to get an eye exam?<br>Were any factors more important than others? |
| **Knowledge-creating Experiences**<br>How did you learn about eye exams?<br>How did you know you needed one?<br>What factors helped you to know?<br>At the time that you were diagnosed with diabetes, what, if anything, were you told about eye care?<br>How did you learn that diabetes can affect your eyes?<br>Have you ever been told by a healthcare provider that diabetes can affect your eyes? |
| **In-Clinic Experience**<br>Can you tell me about your experience the last time you had an eye exam where your pupils were dilated?<br>Can you walk me through your experience? Start with the events leading up to the exam.<br>What was the experience like? What did you think then?<br>Did you encounter problems at any point?<br>What would have made your experience better?<br>How would you describe your relationship with your eye doctor? |
| **Vision Status**<br>When was the first time you noticed changes in your vision?<br>What about your vision has changed recently?<br>What changes startled or concerned you the most? |
| **Competing Concerns**<br>Have you ever cancelled or not shown up to an appointment? If so, why?<br>If you can remember, what happened?<br>What would make you reschedule or cancel an eye exam? |
| **Emotional Context**<br>When was the last time you had a dilated eye exam?<br>Tell me about how often you get an eye exam.<br>How often do you think you should have an eye exam?<br>If there's a difference between those two answers, why?<br>How did making an appointment make you feel?<br>How did getting an eye exam make you feel?<br>What would you have changed about the experience of getting an eye exam? |

We transcribed all interviews for analysis. We applied grounded theory and the constant comparative method to analyze the transcripts [9]. We reviewed the transcripts line by line and referenced the codebook from our previous study to identify new themes or variations [8]. We contextualized health behaviors using individual, interpersonal, and institutional/structural factors based on the socio-ecological model (SEM). We conducted the analysis using NVivo software, version 12 (Melbourne, Australia) and Dedoose, version 9.0 (Los Angeles, CA). To further enrich the analysis, we recruited and trained a community partner (A.N.) to assist with the coding and analysis of the eight interview transcripts. The study authors (K.N., J.A., and A.N.) held two 45-minute discussion sessions to ensure depth of the analysis and incorporation of insights from all team members. We did not return transcripts to participants for feedback or corrections, but participants were provided with a slide deck summarizing the study’s findings and were invited to offer feedback during the interview.

## Results

We conducted interviews with four participants in the severe disease cohort and four participants in the unengaged cohort to reach thematic saturation. The severe disease cohort was 75% male with an average age of 58.5 years (range 52-64). The unengaged cohort was 50% male with an average age of 61.8 (54-71). Participants in the severe disease cohort were diagnosed with diabetes on average 24.9 years ago (range 3-36), while those in the unengaged cohort were diagnosed on average 7.5 years ago (range 4-10). The average time since the last eye exam was 109.3 days (range 6-365) for the severe disease cohort, and 840.5 days (range 366-1825) for the unengaged cohort. Additional participant demographic details are provided in Table 2.

**Table 2.** Participant demographic information.

|  | Original Cohort [8] | Severe Disease Cohort | Unengaged Cohort |
| --- | --- | --- | --- |
|  | n = 24 | n = 4 | n = 4 |
| **Age, average (range)** | 57.7 (44-73) | 58.5 (52-64) | 61.8 (54-71) |
| **Gender, No. (%)** |  |  |  |
| Male | 14 (58) | 3 (75) | 2 (50) |
| Female | 10 (42) | 1 (25) | 2 (50) |
| **Race, No. (%)** |  |  |  |
| Black/African American | 14 (58) | 3 (75) | 4 (100) |
| White | 4 (17) | 0 | 0 |
| More than 1 race | 0 | 1 (25) | 0 |
| Other/No answer | 6 (25) | 0 | 0 |
| **Ethnicity, No. (%)** |  |  |  |
| Hispanic/Latino/a/x | 3 (13) | 1 (25) | 0 |
| Non-Hispanic/Latino/a/x | 21 (88) | 3 (75) | 4 (100.0) |
| **Insurance Status, No. (%)** |  |  |  |
| Insured | 23 (96) | 4 (100) | 4 (100) |
| Uninsured | 1 (4) | 0 | 0 |
| **Eye Exam Frequency, No. (%)** |  |  |  |
| Annually or more frequently | 17 (71) | 4 (100) | 0 |
| Biennially or less frequently | 7 (29) | 0 | 4 (100) |
| **Duration of diabetes, No. (%)** |  |  |  |
| 5 years or less | 8 (33) | 1 (25) | 1 (25) |
| 10 years or less | 6 (25) | 0 | 3 (75) |
| More than 10 years | 10 (42) | 3 (75) | 0 |

In the severe disease cohort, eight recurring themes emerged: vision status, emotional context, competing concerns, resource availability, cues to action, knowledge-creating experiences, in-clinic experiences, and patient-doctor relationship (Fig 1). In the unengaged cohort, seven themes emerged: vision status, emotional context, competing concerns, resource availability, cues to action, knowledge-creating experiences, and in-clinic experiences (Fig 2). All themes identified in the severe disease cohort, excluding the patient-doctor relationship, were also found in the unengaged cohort. Notably, the patient-doctor relationship theme was unique to the severe disease cohort. Many themes identified in this study were also present in the framework previously developed by our research group [8].

**Fig 1.**
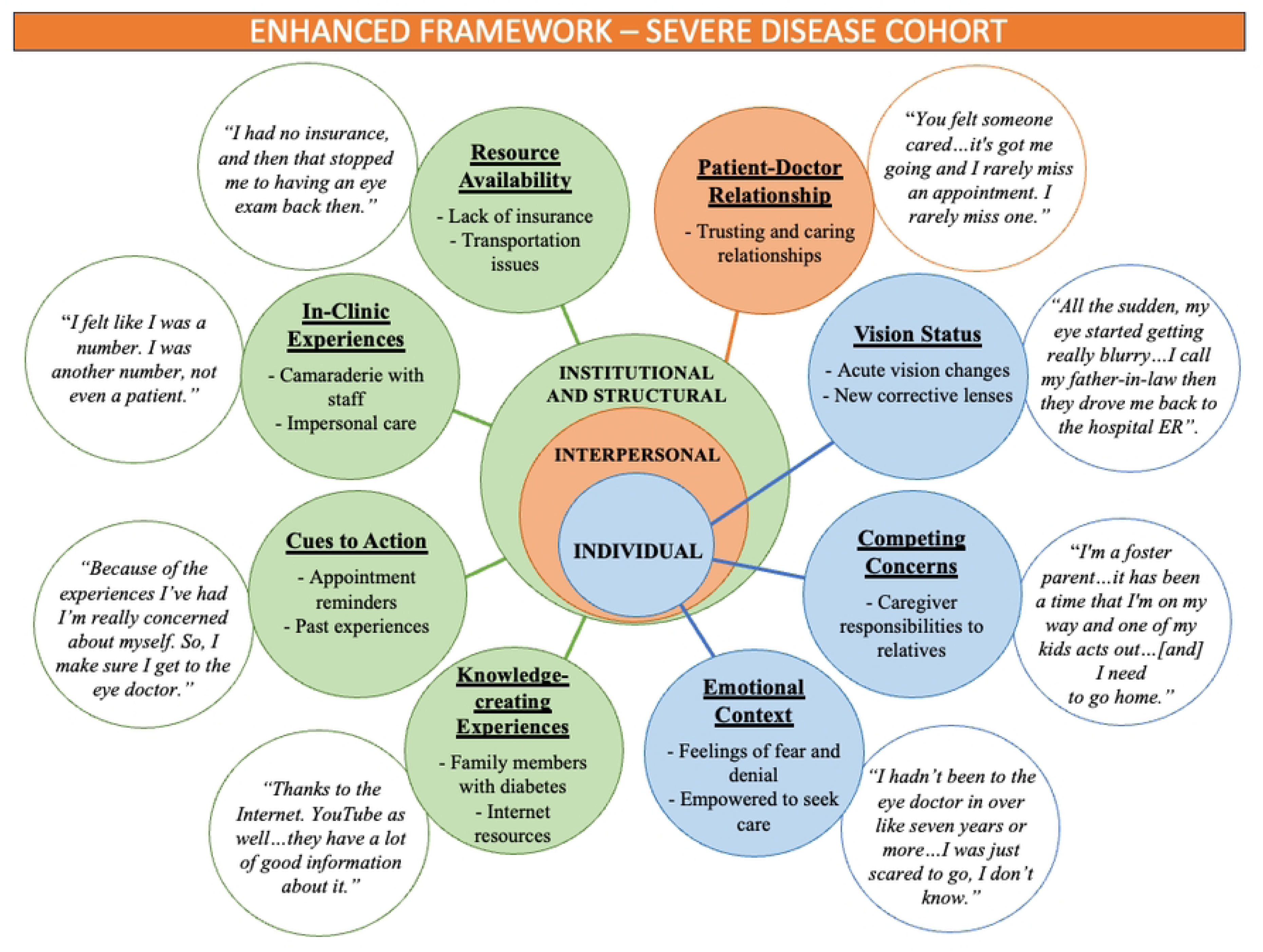
Theoretical framework of themes affecting DR screening adherence in a severe disease population.

**Fig 2.**
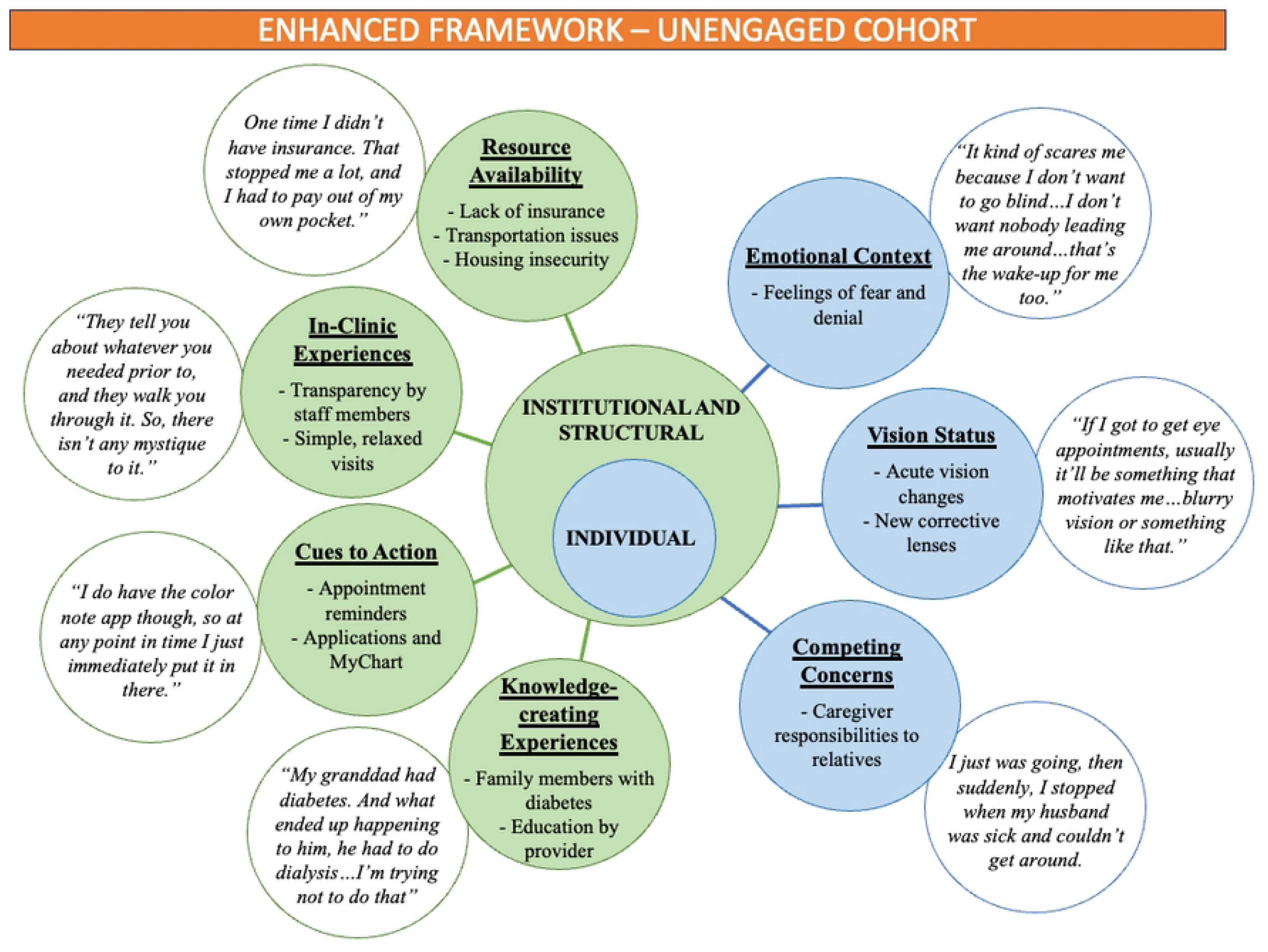
Theoretical framework of themes affecting DR screening adherence in a population not engaged with eye care.

We classified these themes into three levels based on the SEM: 1) Individual level, 2) Interpersonal level, and 3) Institutional and Structural level. As in our previous study [8], the individual level includes vision status, competing concerns, and emotional context. The institutional and structural level includes resource availability, cues to action, knowledge-creating experiences, and in-clinic experiences. The interpersonal level includes the patient-doctor relationship, which was newly identified in this study.

### 1) Individual Level

#### Vision Status

Participants reported that changes in vision status influenced their adherence to DR screening. Both cohorts reported that the need for new glasses often motivated them to seek eye exams. One participant from the severe disease cohort explained: “I’m just trying to keep up with new frames.” Similarly, a participant in the unengaged cohort said: “I need to change my eyeglasses, because the ones I have, I’ve had those [for] about eight years.”

Acute vision loss also prompted participants to seek eye care. A participant from the severe disease cohort described how a sudden change in vision led to an emergency eye exam: “All of a sudden, my eye started getting really blurry…I called my father-in-law, [and] they went and picked me up, then they drove me back to the hospital emergency room.” Other participants in this cohort sought care due to symptoms like twitching, seeing blood vessels in their eyes, or experiencing transient blurry vision. One participant in the unengaged cohort was prompted to seek care after noticing a shadow in her vision: “I could see a shadow coming around my eye. When I got it, I called for Yale.”

Participants in both cohorts did not perceive a need for eye screening if their vision remained stable. A participant in the severe disease cohort stated: “I did not pay so much attention to that because I have 20/20 vision…I was not rushed or worried to go back to the treatment, because like I said, I did not understand how bad my eye was.” Similarly, a participant from the unengaged cohort remarked: “If I got to get eye appointments, then usually it’ll be something that motivates me to…blurry vision or something like that.”

#### Competing Concerns

Both cohorts reported that competing concerns prevented them from accessing eye screening. A participant in the severe disease cohort explained: “I’m a foster parent. And, you know, it has been a time that I’m on my way and one of my kids acts out, you know, and so I’m in the car [and] I need to go home.” Similarly, a participant in the unengaged cohort said: “I just was going, then suddenly, I stopped when my husband was sick and couldn’t get around. I went when I could, but if I had to take him or let him go to the doctor or something, I didn’t have a way.”

#### Emotional Context

Fear and denial influenced screening adherence in both cohorts. One participant in the severe disease cohort reflected on their long delay seeking care due to fear: “I hadn’t been to the eye doctor in over like seven years or more. I was just scared to go, I don’t know. I don’t know why. I guess I didn’t want to know what was going on with my eye.” Similarly, a participant from the unengaged cohort said: “It kind of scares me a little bit because I don’t want to go blind. I want to see. I don’t want nobody leading me around, you know…that’s the wake-up for me too.”

Other participants in the severe disease cohort accepted their condition and were empowered to seek care. One participant stated: “Well, it really didn’t bother me. It takes a lot to really get me riled up about anything. I took care of it. It was rectified. And I know what to do.” Another participant added: “I feel right now at this moment my vision is stable, that it is controlled. I’m not afraid to be blind at this point. Honest to you. Like before, I was afraid to be blind.”

### 2) Interpersonal Level

#### Patient-doctor relationship

Relationships with healthcare providers played a significant role in promoting screening adherence for participants in the severe disease cohort. Positive interactions with physicians fostered trust and encouraged continued engagement in care. One participant remarked: “You felt someone cared…that’s what brought me home and it’s got me going and I rarely miss an appointment. I rarely miss one.” Another participant noted that their doctor’s caring attitude motivated them to take better care of their health: “She had one of the best attitudes I have ever, ever, ever run across, you know…because by her care, and showing she cared, made me honestly start caring about me.”

In contrast, participants in the unengaged cohort did not identify the patient-doctor relationship as a major factor in their eye care utilization.

### 3) Institutional and Structural Level

#### Resource Availability

Participants in the unengaged cohort frequently reported lack of insurance as a major barrier to accessing eye care. One participant said: “One time I didn’t have insurance. That stopped me a lot, and I had to pay out of my own pocket.” Another participant echoed this sentiment: “I didn’t have the insurance at that time too…I had to pay for everything out of pocket.” Lack of transportation also emerged as a significant barrier. One participant said: “Well, just I wasn’t able to get there, that’s all. If I would have had somebody to come bring me, I would have been there. And then I have to go through the stress of asking somebody to bring you, have to deal with their attitude and whatever they were supposed to be doing at the time. I don’t want to go through that. That’s why I didn’t make it.”

Housing insecurity also impacted access to care for some participants. One participant shared: “I’m homeless as we speak but I have somewhere to be at for now. But I am definitely homeless.” When asked how housing insecurity affected eye care, they said: “It’s difficult, but I manage.”

Similarly, participants in the severe disease cohort reported that lack of insurance and transportation prevented them from accessing care. One participant explained: “I had no insurance, and then that stopped me from having an eye exam back then. And then from there to ’18, that’s when the problems occurred in my eye.” Another participant said: “I think if I missed an appointment, it was because I had no one to drive me down there.”

#### In-clinic Experiences

Participants in both cohorts described how in-clinic experiences influenced their adherence to screening. In the severe disease cohort, some participants highlighted positive interactions with staff. One participant shared: “We hit it off. And the first time I walked through the door, we had, you know, me, her, her assistants, and the nursing staff. Very good. Nothing bad to say about any of them.” However, participants in this cohort also reported negative experiences. One participant expressed feeling dehumanized: “I felt like I was a number. I was another number, not even a patient.” Another participant echoed this sentiment, emphasizing a desire for more personal treatment: “You wanted to be treated as a person. So, that’s the only problem I had.”

Similarly, participants in the unengaged cohort described how in-clinic experiences affected their use of eye care. One participant noted: “When I go there, everybody remembers me.” Another participant appreciated the clear guidance provided during visits: “They’re very informative. They tell you about whatever you needed prior to, and they walk you through it. So, there isn’t any mystique to it.”

#### Cues to Action

Both cohorts identified various cues that prompted them to seek eye care. These included reminders from office staff and mobile apps. One participant in the severe disease cohort shared: “You know, they call you a day or so ahead of time and let you know, you have an appointment.” Similarly, a participant in the unengaged cohort said: “I do have the color note app though, so at any point in time I just immediately put it in there.” For some participants, personal experiences were a strong motivator to prioritize eye care. One participant in the severe disease cohort said: “Because of the experiences I’ve had with myself, and I’m really concerned about myself. So, I make sure I get to the eye doctor.”

#### Knowledge-creating Experiences

Participants in both cohorts reported that learning about diabetes motivated them to seek eye care. Many learned from and shared knowledge with family members. One participant in the severe disease cohort said: “But after that, learning how to control the pancreas, the fluid, how to control myself in food…I’m the expert. My sisters, I’m helping them when their diabetes is out of control.” Similarly, one participant in the unengaged cohort said: “My granddad had diabetes.

And what ended up happening to him, he had to do dialysis, at the end of the day. I’m trying not to do that.”

Other participants sought knowledge from diverse sources. One participant in the severe disease cohort credited online resources: “Thanks to Internet. YouTube as well…they have a lot of good information about it. College, universities, they have a real fact and education.” Another participant in the unengaged cohort received information on diabetes from their primary care physician: “I had spoken to my primary care and then I read all the pamphlets that they give you.”

## Discussion

Annual eye exams are recommended for early detection of diabetic eye diseases as DR often progresses without noticeable symptoms in the early stages [10]. This study builds upon and validates a framework previously developed by our group to identify barriers to and facilitators of DR screening among individuals with severe disease and those unengaged in eye care [8]. In addition to the themes identified in our previous study, this study emphasizes the critical role of the patient-doctor relationship. It also incorporates the unique perspectives of individuals who are not engaged in the eye care system.

A common theme across both cohorts was how acute changes in vision served as a primary motivator to seek eye care. Participants often did not perceive a need for screening if their vision remained stable. This is consistent with our previous study, which identified acute vision changes and the need for new glasses as strong motivators for seeking eye care [8]. Similarly, Chou et al. found that “no need” was one of the most frequently cited reasons for not obtaining DR screening within the past year [11]. This reinforces the importance of addressing patient perceptions of need as a critical factor in promoting routine DR screening.

The perceived lack of need for eye care when vision remains adequate highlights a significant gap in our education to patients about DR. Routine DR screenings should be done prior to symptom onset since DR is often asymptomatic in the early stages. The onus of patient education should not be on the patients themselves, but instead is the responsibility of individual providers and the healthcare system. When we asked participants where they learned about diabetic eye disease and screenings from, few cited the healthcare system. Most reported relying on family members or the internet for information. This is consistent with our previous study, where few participants mentioned learning that diabetes could affect vision from their healthcare providers [8]. Similarly, Nwanyanwu et al found that 10.6% of US adults aged ≥40 (9.8 million individuals) were unaware of their DR diagnosis and 70.1% of individuals who demonstrated photographic evidence of retinopathy were unaware of their condition [12]. These results align with many other studies [13–18], which suggests a substantial need for the healthcare system and individual providers to improve communication about the importance of DR screening to patients before they experience visual changes.

Resource availability emerged as a strong theme influencing screening access among participants in the unengaged cohort. Participants cited transportation issues and lack of insurance as the most significant barriers. Transportation issues were particularly burdensome because participants were reluctant to rely on others for assistance. Similarly, lack of insurance was a critical obstacle, as it required many participants to pay out of pocket for eye care. Furthermore, some participants expressed uncertainty about whether their existing insurance would cover DR screening. These findings align with previous studies, which highlight cost-related delays and difficulties traveling to appointments as common barriers to eye care [7,11,19–22]. Participants in the severe disease cohort also reported insurance and transportation issues, though to a lesser extent than the unengaged cohort. Collectively, these results suggest that addressing insurance and transportation issues through targeted interventions such as patient navigation could enhance access to DR screening.

Interestingly, participants in our prior study did not identify transportation issues as a significant barrier to DR screening despite participants in both studies being from relatively the same geographical area [8]. This discrepancy may be attributed to differences in recruitment settings. We recruited participants in our previous study in a location with co-located ophthalmology and primary care community-based clinics, which likely reduced the impact of transportation barriers. This highlights how the organization of healthcare services can influence the accessibility of care.

Notably, participants in the severe disease cohort reported long-term, trusting relationships with their physicians that fostered ongoing engagement in eye care. Many participants emphasized that these relationships were vital in empowering them to take charge of their health. In contrast, participants in the unengaged cohort did not report similar relationships. This difference suggests that positive patient-doctor relationships may play a critical role in promoting DR screening adherence. This is also supported by other studies, which have shown that high-quality communication and cultural humility in patient-doctor interactions can significantly increase screening adherence [23]. Similarly, a lack of trust in providers is a barrier to eye care shown in other studies [13].

To improve DR screening access, innovative strategies must be implemented to effectively communicate information, build positive patient-doctor relationships, and address resource barriers. Clinical systems should prioritize educating patients about the link between diabetes and vision loss and emphasize the importance of routine screening. This can be achieved through clinician-reviewed or developed educational videos, which have been shown to improve patient understanding and promote long-term adherence to treatment [24,25]. Other strategies to improve education about DR include patient-centered communication techniques, such as asking open-ended questions, expressing empathy, and sitting at eye-level, which can not only aid in effectively communicating information but are also essential in building positive patient-doctor relationships [26–28]. Strengthening this relationship is key to improving DR screening adherence and can also be achieved by measures such as mitigating time limitations during appointments [29], mutual agreement and valuing of patient goals [30], and improving patient perception of clinician expertise [31,32].

To address resource barriers, telemedicine, which expanded significantly during the COVID-19 pandemic, can partially alleviate transportation issues by allowing patients to access care remotely [33]. Expanding awareness and access to healthcare mobility services, such as ride-sourcing options, is another promising strategy [34,35]. We can also use community-based participatory research to simultaneously identify barriers to care and develop targeted interventions to address these barriers. For example, Frimpong et al previously used community-based focus groups to design a culturally relevant and practical digital health tool for individuals with diabetes based on the needs of people in these focus groups [36].

When implementing screening protocols, it is also imperative to establish reliable follow-up pathways to ensure timely management and treatment of sight-threatening conditions. Many of the barriers to DR screening identified in this study, such as medical comorbidities, lack of a consistent healthcare provider, and socioeconomic factors, are also known to affect adherence to follow-up care [37–41]. Addressing these interconnected barriers is critical for improving long-term adherence to screening.

The strengths of this study include the qualitative approach, which allows for an in-depth and nuanced exploration of participants’ experiences. However, this study also contains limitations. Generalizability to broader populations may be limited due to a small number of participants being interviewed in a single geographic area. This study also has potential for researcher bias, as question framing and interactions with participants may potentially influence responses. Additionally, social desirability and response bias may influence this study as participants may modify their responses to align with perceived social norms or expectations.

Further research is needed to better understand DR screening adherence and develop interventions to overcome potential barriers. This study expanded our current understanding by evaluating the perspectives of individuals with severe disease as well as those unengaged in eye care. This allowed for a deeper understanding of the importance of improving education about DR, strengthening patient-doctor relationships, and mitigating transportation and insurance issues. The groups of participants evaluated in this study are particularly vulnerable to vision loss and therefore, it is essential to both understand their relationship with DR screening and develop strategies to enhance their engagement with eye care.

## Data Availability

All relevant data are within the manuscript and its Supporting Information files.

## Acknowledgements

We would like to thank all the participants for their time and openness in sharing their experiences with us and our community partners for their valuable insights.

---

**S1 Appendix. Interview slide deck**

## Notes

### Competing Interest Statement

The authors have declared no competing interest.

### Author Declarations

The institutional review board (IRB) of Yale University approved this study (HIC 2000029860).

